# Development and deployment of a digital platform for the collection of consistent non-communicable disease epidemiological data across multiple low and middle-income countries: A user-centred design approach

**DOI:** 10.64898/2026.08.06.26359759

**Authors:** Wubin Xie, Ananya Gupta, Mokbul Hossain, Mehedi Hasan, Soren Brage, Nita Forouhi, Ashutosh Yadav, Vindya Rajakaruna, Manoja Gamage, Sara Mahmood, Rajendra Pradeepa, Vinita Jha, Anuradhani Kasturiratne, Prasad Katulanda, Khadija Irfan Khawaja, Malay Kanti Mridha, Fred Hersch, Ranjit Mohan Anjana, John C. Chambers, Ian Y. Goon

## Abstract

**Background:** A critical challenge for large-scale multi-country population health studies is the ability to collect consistent data across many sites and time periods and ensure that the data collected are valid, complete and comparable. To address these challenges, we developed a fit-for-purpose digital data collection platform using mobile devices, to enable creation of the South Asia Biobank study.

**Objective:** To describe the process by which a digital platform was designed, developed and deployed across four countries in South Asia; to demonstrate how the platform enabled field research teams located across these countries to collect non-communicable diseases epidemiological data consistently.

**Methods:** A user-centred design approach was employed for the development of the digital platform to address both the diversity and dynamic nature of study requirements. This approach uses 5-step iterative loops that, with each iteration, produce a usable prototype version of the software that was then tested by potential users of the platform. Qualitative interviews and quantitative system usability assessments were conducted, and findings utilised as input for the start of the next iterative loop. Once the process was completed, a working version of the software was developed for the use in the study.

**Results:** Over the course of four iterative loops, the platform was progressively built and tested to ensure its functionality met the requirements of the study. Detailed feedback was collected from key stakeholders and incorporated into the platform with each new version of the applications. The platform leverages advances in mobile and medical device technology along with software integration capabilities to enable efficient and consistent data collection, along with the ability to review data quality and make improvements to the data collection process in real-time. The successful deployment of the data platform has enabled collection of comprehensive baseline data from 205,536 participants in four South Asian countries.

**Conclusions:** Using user-centred design principles, it is possible to develop and deploy a comprehensive digital surveillance data management platform that allows consistent and high-quality data collection in population health studies in remote settings. To the best of our knowledge, this is the first platform that enables the integrated capture of health assessment data from a wide variety of medical equipment that is tailored for deployment in a range of LMIC settings.

## Introduction

Type-2 diabetes mellitus (T2DM) and cardiovascular disease (CVD) are leading and closely interlinked global health challenges [1]. In South Asia, the most populous and most densely populated geographical region in the world, the burden of T2DM and CVD are especially high [2]. In recent years, the prevalence of diabetes in South Asia has risen more rapidly than in other large geographic regions of the world [3]. Identifying the major risk factors for T2DM and CVD provides essential insights into disease aetiology and facilitates the development of effective approaches for prevention and treatment [4]. Despite representing a large proportion of the global population, South Asians are not well represented in large-scale longitudinal studies.

To address this gap and better understand the wide range of exposures that contribute to developing T2DM and CVD in South Asians, a unique longitudinal population study, the South Asia Biobank (SAB), has been established. The SAB was launched in 2018 as a partnership between collaborating centres in Bangladesh, India, Pakistan, Sri Lanka and the U.K. [5]. A key component of the SAB is to establish a network of non-communicable disease (NCD) surveillance sites in each of the member countries, using common protocols and platforms to complete structured health assessments on a representative sample of 200,000 South Asians aged 18 years and above residing at surveillance sites [5]. The study aims to collect information on demographic, lifestyle, clinical, environmental, genomic variables, as well as information on subsequent health outcomes.

A critical challenge for population health studies such as the SAB is the need for consistency of data collected across numerous sites and time periods to ensure data collected are valid and comparable [6,7]. The multi-national nature of the SAB study, with each country having multiple surveillance sites with varying characteristics (e.g. rural and urban), poses a unique challenge for ensuring the consistency of data collected [8]. In addition, intermittent internet connectivity and the need for surveillance teams to move regularly from one surveillance site to another, present additional challenges [9]. Digital platforms that are portable and configurable have the potential to address these challenges and support high-quality data collection [10–13]. The overarching design requirements for building a platform for the SAB study were that it had to (i) operate without internet connectivity, (ii) be readily scaled across multiple countries, (iii) minimise human data entry via integrations with screening devices and (iv) developed with open-source frameworks to ensure the platform can be more readily updated and maintained. An assessment of the available options on the market did not yield any system which satisfied the overarching requirements. We decided to develop a fit-for-purpose digital data collection and management tool for deployment across the study sites to strengthen the ability of the research teams from different countries and organisations to collect consistent and high-quality data.

In this manuscript, we describe the process by which a digital platform was designed, developed and deployed across four countries in South Asia and demonstrate how it has enabled teams located across the South Asian region, to collect consistent comparable data for analysis. The modular, robust and flexible platform developed for use in the SAB study can be a valuable addition to the Global Health research community looking to leverage modern digital technologies for consistent health assessment and surveillance data collection. It is crucial that these platforms are well-designed and are made available for the benefit of the community and ecosystem.

## Methods

The SAB study involves seven partner organisations conducting protocol driven health measurements and questionnaires across five distinct regional hubs in four countries. Given the cultural and geographic diversity between regional hubs and settings (e.g. rural vs urban, different regulations), novel and localised health screening formats were required to reach local populations and conduct NCD surveillance health assessments [5]. The assessment process is organised into 13 separate screening stations, with over 1,000 data points collected from each participant using the corresponding equipment or questionnaire tool (**Table 1**).

**Table 1.** Summary of data to captured in digital data collection platform.

| Data collected | Screening equipment / tool |
| --- | --- |
| Enumeration and invitation | Internally developed questionnaire |
| Registration | Internally developed questionnaire |
| Height | SECA 213 |
| Body composition (including weight, body fat percentage, skeletal muscle percentage, visceral fat level) | OMRON BF-511 |
| Blood pressure | OMRON HEM-9210T |
| Electrocardiogram (ECG) | GE MAC2000 |
| Lung function test (Spirometry) | Nuvoair Air Next |
| Retinal imaging (Fundoscopy) | Crystalvue NFC-700 |
| Diet assessment (multiple-pass 24-hour recall) | Intake24 digital dietary recall system |
| Physical activity (7-day physical activity via accelerometry) | Axivity AX3 |
| Biological sample collection and point of care blood tests (Glucose and Total cholesterol) | Jana Care Aina mini (Glucose)<br>Jana Care Aina (Total Cholesterol) |
| Health and lifestyle questionnaire | Internally developed questionnaire |
| Health assessment report (incorporating CVD risk score) | Custom algorithm based on WHO Hearts leveraging Open Health Algorithm service |

The screening formats considered included performing assessments in fixed locations such as a clinic, in mobile health vehicles which were transported to the site daily, or hybrid formats where selected screening activities were carried out in participant’s households (see supplementary Figure 1 for study site map). As the screening format protocol definition and digital platform development proceeded in parallel, a software development approach that was able to account for changing requirements and yet deliver a platform that was robust and flexible enough to cater for a range of users and settings was required. A study technology team comprising members with experience in healthcare research and technology development was formed to lead technology development, supported by a technology working group consisting of members from each region.

A software development process incorporating user-centred design (UCD) methodology was selected to build the platform. UCD is an iterative software development methodology in which the development team focuses on the users and their needs in each phase of the development process [14]. Widely employed in modern software development [15–17], the iterative and user-focused nature of the UCD method ensures that the software platform built caters to the needs of the research data collector’s workflow and supports consistent and high-quality data collection. The multi-disciplinary technology development team consisting of domain experts, software engineers and user design researchers utilised UCD in the development of the digital platform for the SAB and involved the use of 5-step loops that with each iteration, produced a usable version of the software that was increasingly aligned with the needs of its users (research data collectors and managers). The process was completed when a sufficiently developed version of the software was produced that could be deployed for use in the study.

The 5-step UCD loop, summarised in Figure 1, involved firstly discussions with the study investigators and research managers to define the requirements of the prototype to be built in terms of functionality and user experience, given the study protocol (*Define*). This was followed by interviewing field data collectors and managers to understand their workflow, needs and challenges when carrying out their given data collection tasks (*Empathise*). The software development team then undertook a process of ideation whereby designers and software engineers shared and refined ideas for how the prototype was to be built (*Ideate*). Once the designs and parameters were agreed upon, a usable prototype of the software with the required features was built (*Prototype*). The final step was to allow users to test the prototype by simulating its use in real-world settings (*Test*). During the process of testing the prototypes, insights on the usability and functionality of the platform were collected from a group of ten research assistants, with experience in field data collection for similar studies. These insights included quantitative usability assessments captured via the System Usability Scale tool [18,19], and qualitative feedback via interviews with users. The insights and feedback collected at the end of each iterative loop then serve as the starting point for the next iteration.

**Figure 1.**
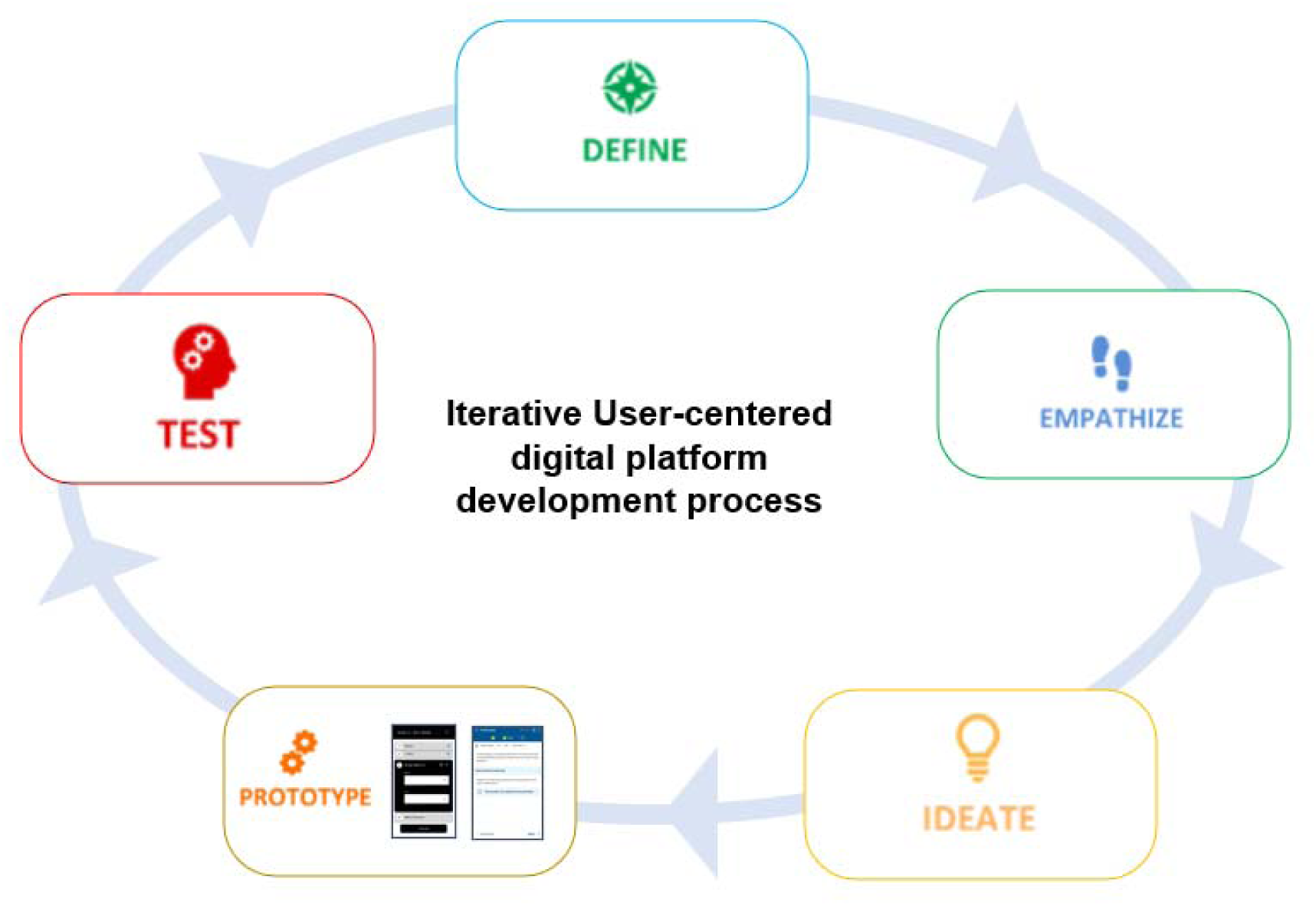
User-centred design iterative methodology utilised in platform development.

The digital platform was developed across four iterative loops, with approximately 40 days required to complete each loop, and with a testable version of the software platform developed progressively through each of the loops. The final iterative loop consisted of a pilot evaluation of the platform in real-world settings. The activities, evaluations performed and development progress within each loop is summarised in **Table 2**. System usability scale (SUS) assessments were conducted on groups of 10 users after the 2nd to 4th iterative loops. An ‘above average’ SUS score of 80 combined with successful completion of the pilot surveillance exercise was deemed as sufficient for commencing deployment in all surveillance sites [20,21].

**Table 2.**
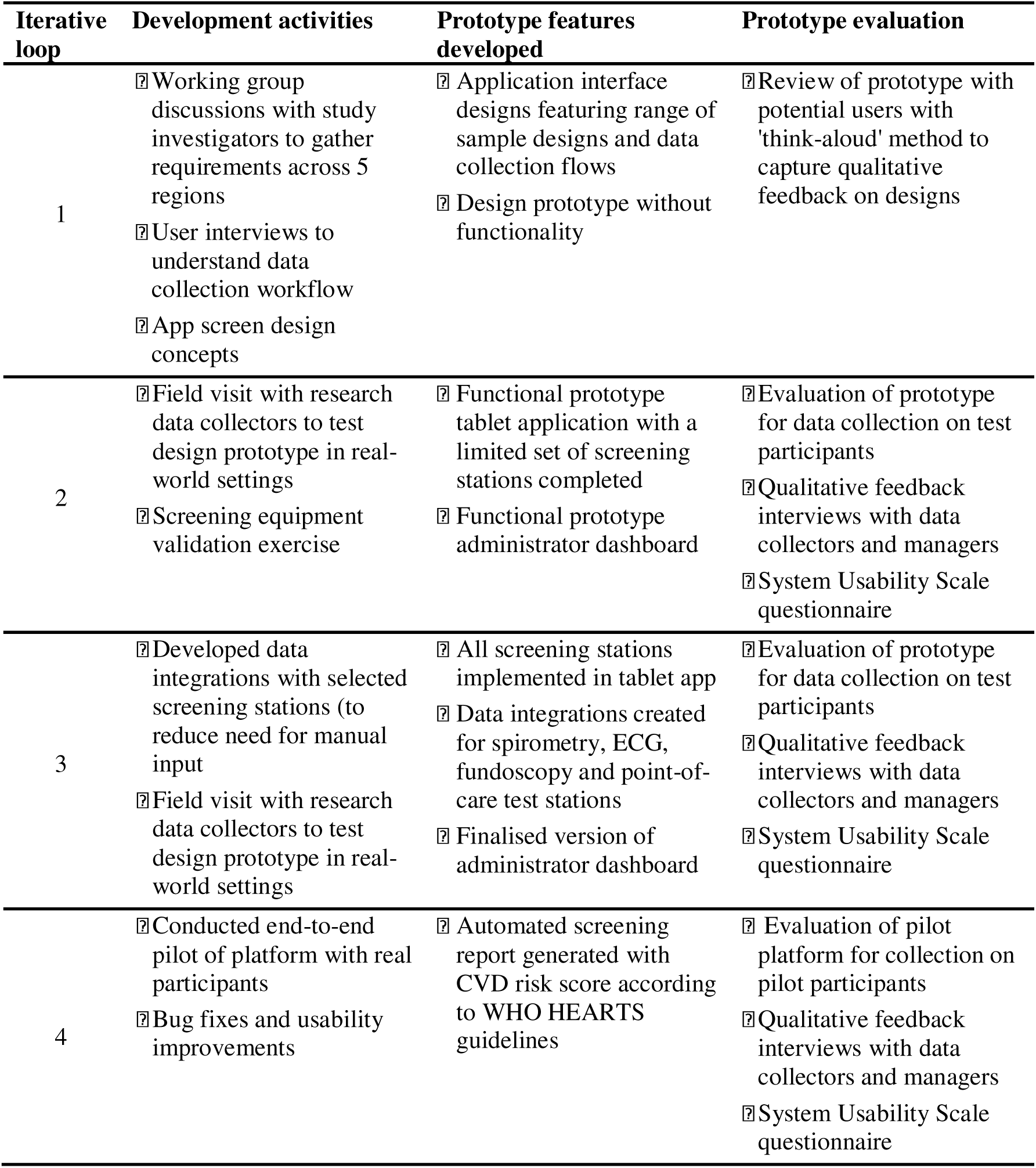
Summary of activities, features and evaluations completed through each iterative loop.

After the technology platform solution was deemed to be sufficiently refined and ready for use in the field, a deployment plan was developed to ensure consistency in the screening operations across the study regions. The deployment plan included a manual of procedures that detailed how surveillance exercises should be conducted, training plans for research team managers and field data collectors, and processes for deploying the platform and screening equipment in the field. After successful deployment of the platform in the field, data quality control checks were implemented to assess data quality and consistency. These checks include data completeness, consistency of the data formats collected.

### Costing approach

To contextualise the resource requirements of the platform, we assembled an indicative costing of its development and deployment from a research-programme perspective. Costs were grouped into three categories: (i) software development, comprising the engineering, design-research and cybersecurity effort across the four user-centred design loops; (ii) recurrent cloud infrastructure, comprising hosting, storage and data transfer on Google Cloud Platform across the region-specific instances; and (iii) training and field deployment, comprising the three-phase training programme, development of the manual of procedures and training materials, and in-field pilot support. Figures were drawn from project financial records and contracts, are expressed in US dollars at 2020 prices, and are reported as approximate values rounded to reflect their indicative nature. Development is reported as a one-off cost, cloud infrastructure as an annual recurrent cost, and training as a one-off cost incurred at launch. The costing excludes the capital cost of screening equipment and tablets, which were procured separately, as well as participant-facing costs, field data-collector salaries, and central study-management overheads, which fall outside the platform itself. The indicative cost per participant was derived by summing one-off development and training costs with cloud infrastructure costs accrued over the 5-year deployment period (2019–2024) and dividing by the 205,536 participants screened. Because these figures reflect the specific labour markets, partnership arrangements and in-kind contributions of the SAB, they should be interpreted as illustrative rather than as a formal economic evaluation.

### Ethical Considerations

The South Asia Biobank is conducted in accordance with the recommendations for physicians involved in research on human subjects, adopted by the 18th World Medical Assembly, Helsinki, 1964, and later revisions. Research approval was obtained from the Imperial College London Research Ethics Committee (reference: 18IC4698) and local institutional review boards in each of the participating countries. Written informed consent was obtained from all participants prior to enrolment and the conduct of any study procedures.

## Results

### Platform development and evaluation

Over the course of four iterative loops, platform prototypes were built and tested to ensure they met functionality requirements of the study. Detailed feedback was collected from key stakeholders and incorporated into the platform with each new version of the applications. Tables 2 and 3 summarise the key requirements and feedback collected during testing, along with the platform features developed in response for each iteration. It took approximately 5 months in total to develop a platform that was ready for pilot testing with surveillance study participants. SUS assessments results for the 2nd to 4th iterative loops from 10 users (mean[SD] age 24.6 [2.8] years, 60% had an undergraduate degree, 10% with a postgraduate degree) were shown in **Table 4**. A substantial improvement in SUS scores was observed from an initial 58.0 (SD 7.4) for the functional prototype after the 2nd loop to 81.8 (SD 4.4) for the pilot platform after the 4th loop. A pilot deployment of the platform was conducted in Sri Lanka in November 2018 with approximately 500 participants recruited. Having demonstrated success in the pilot surveillance exercise and an acceptable SUS score, the pilot version of the platform was considered ready for deployment in all surveillance sites to create the SAB cohort study.

**Table 3.**
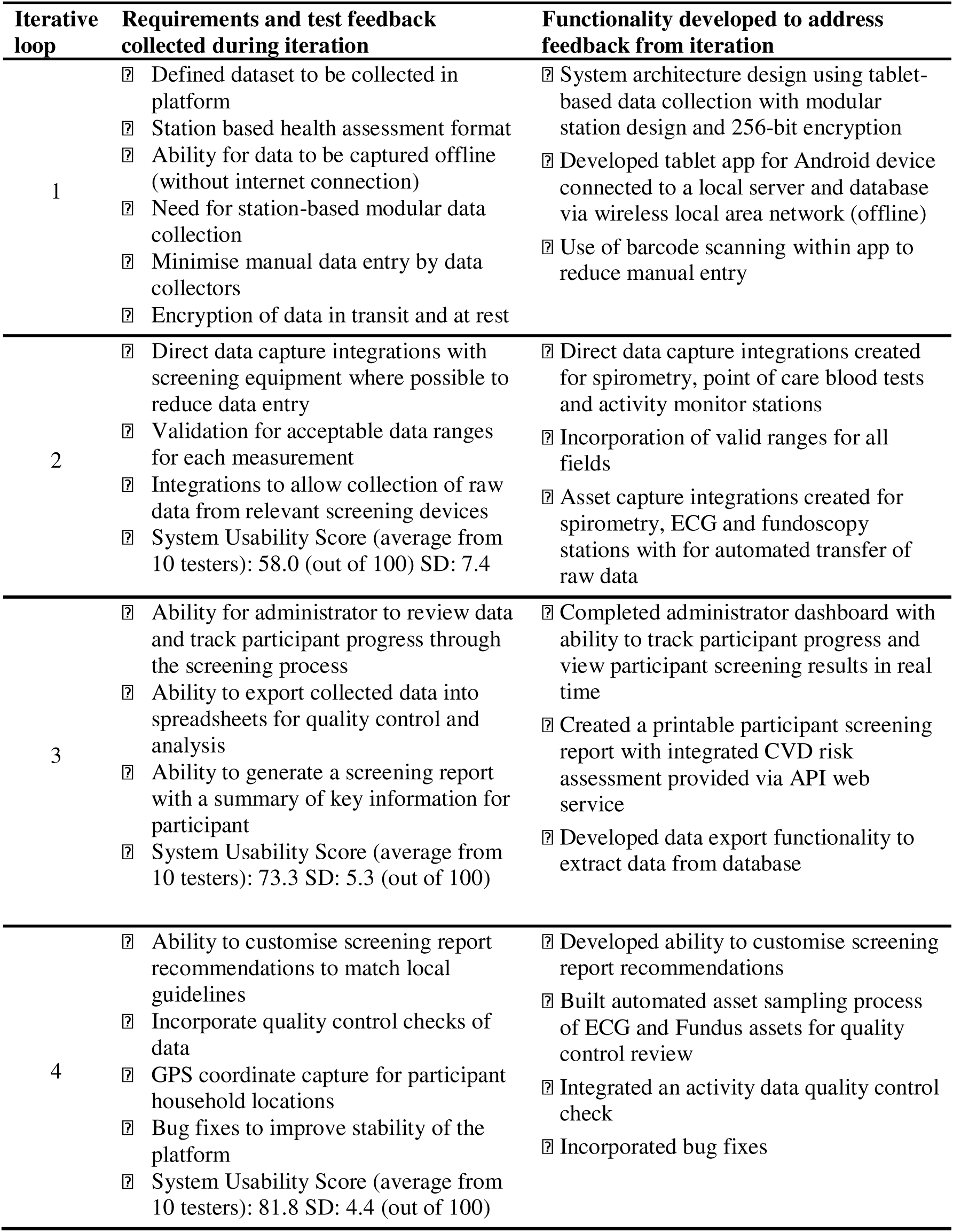
Summary of feedback collected and functionally developed to address feedback.

**Table 4.** System usability scale (SUS) scores by iterative loops.

| Items * | Iterative Loops |  |  |
| --- | --- | --- | --- |
|  | 2<sup>nd</sup> | 3<sup>rd</sup> | 4<sup>th</sup> |
| 1. I think that I would like to use this application frequently | 3.2 | 3.5 | 3.7 |
| 2. I found the application unnecessarily complex | 2.1 | 2.7 | 3.7 |
| 3. I thought the application was easy to use | 2.0 | 2.8 | 3.3 |
| 4. I think that I would need the support of a technical person to be able to use this application. | 3.2 | 3.6 | 3.8 |
| 5. I found the various functions in this application were well integrated. | 2.1 | 2.5 | 2.7 |
| 6. I thought there was too much inconsistency in this application. | 2.5 | 3.2 | 3.5 |
| 7. I would imagine that most people would learn to use this application very quickly | 2.1 | 2.8 | 2.8 |
| 8. I found the application very cumbersome to use | 2.4 | 3.1 | 3.4 |
| 9. I felt very confident using the application | 1.8 | 2.3 | 2.7 |
| 10. I needed to learn a lot of things before I could get going with this application | 1.8 | 2.8 | 3.1 |
| **System Usability Score** | **58.0** | **73.3** | **81.8** |

\* Items 2, 4, 6, 8, 10 were reverse coded before analysis

### Overview of the platform developed

The platform is based on a classic multi-tier web-based architecture and designed to enable a hub and spoke deployment model in the field where data from each of the participating teams is collected locally in each ‘spoke’ and transmitted to a central ‘hub’ for processing and analysis.

#### Platform architecture

Tablet-based data collection was selected as the primary tool for driving the data collection at each station. The Android OS was chosen as the operating system platform due to the greater flexibility it offers in app development, testing and deployment, along with users’ familiarity due to widespread usage in South Asia and the availability of low-cost tablets. The platform is underpinned by an administrator web-dashboard application built using Laravel, an open-source PHP framework and connected to a database built using PostgreSQL, open-source relational database management system. The use of these open-source components ensures expertise for maintaining and extending the platform is widely available and enhances its scalability as multiple instances can readily be deployed in any required location.

#### Offline data collection

To enable offline data collection, components of the screening platform comprising of tablet devices for data collection, a local server laptop which hosts the database, and equipment for performing the health assessments are connected via a wireless local area network (WLAN). This set up enables use of software application, wireless data collection, transfer and storage without the need for an internet connection (**Figure 2**). Additionally, when coupled with the use of uninterruptible power supply devices, the platform is able to operate for several hours without reliable power. The whole platform including screening equipment can be packed into a one cubic meter space, making it highly portable and deployable even in the most remote locations. This is crucial for enabling consistent data collection across the varied settings in this study.

**Figure 2.**
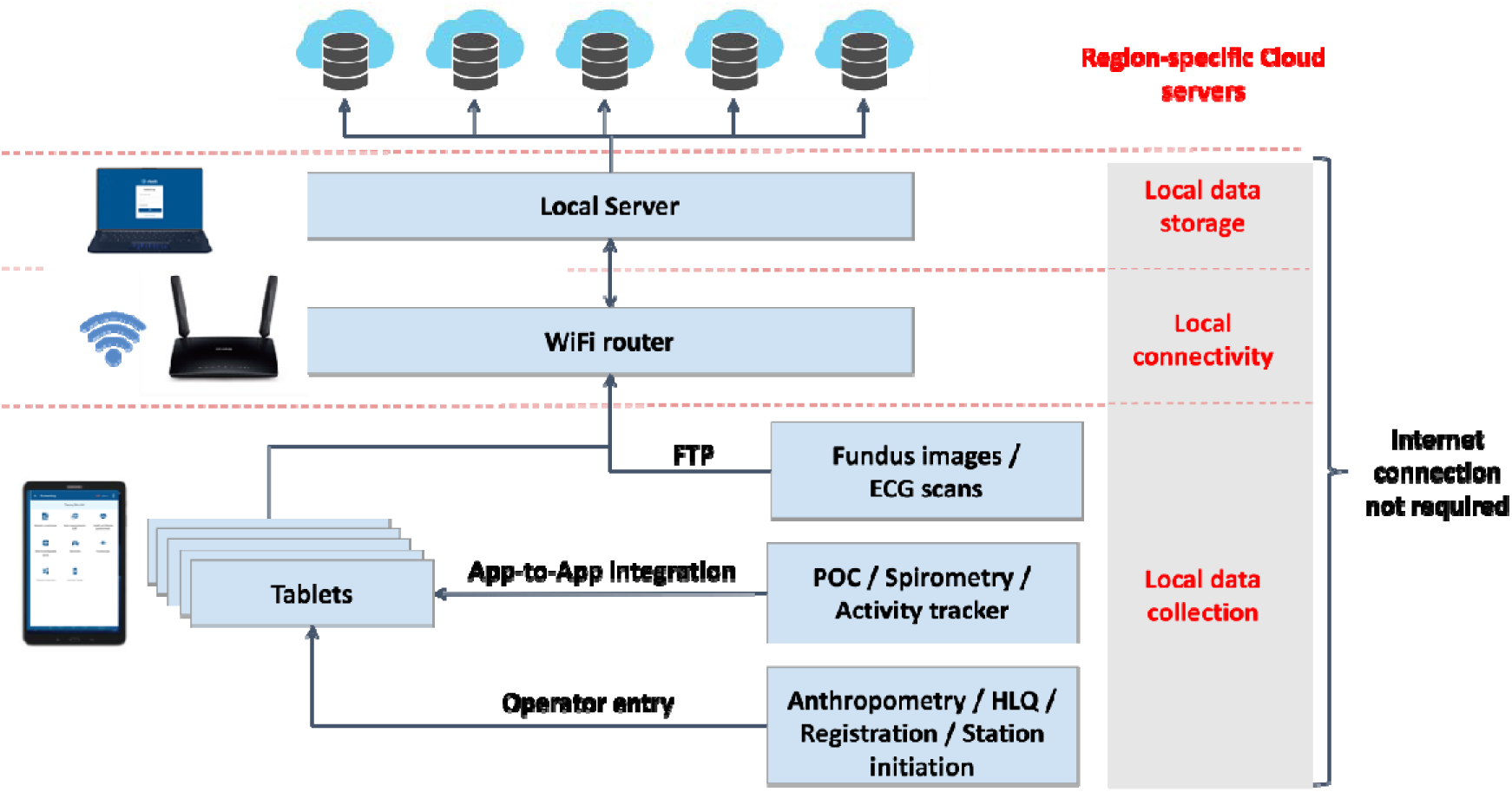
Platform architecture enabling offline data collection and connected device integrations.

#### Modular station-based design

The data collection tablet application was designed with each assessment station as a distinct module to enable for a flexible screening process whereby participants are able to attend any assessment station in any order (**Figure 3**) including in different locations and on different days. The modular design allows for data collectors to move seamlessly from one station to another if required, allowing multiple collectors to perform one station if there is a backlog of participants. This approach also allows for additional stations to be readily integrated into the platform if required. Additionally, specific modules for participant invitation in the community, biological sample management and generating health assessment reports for participants were built to support the end-to-end surveillance study workflow.

**Figure 3.**
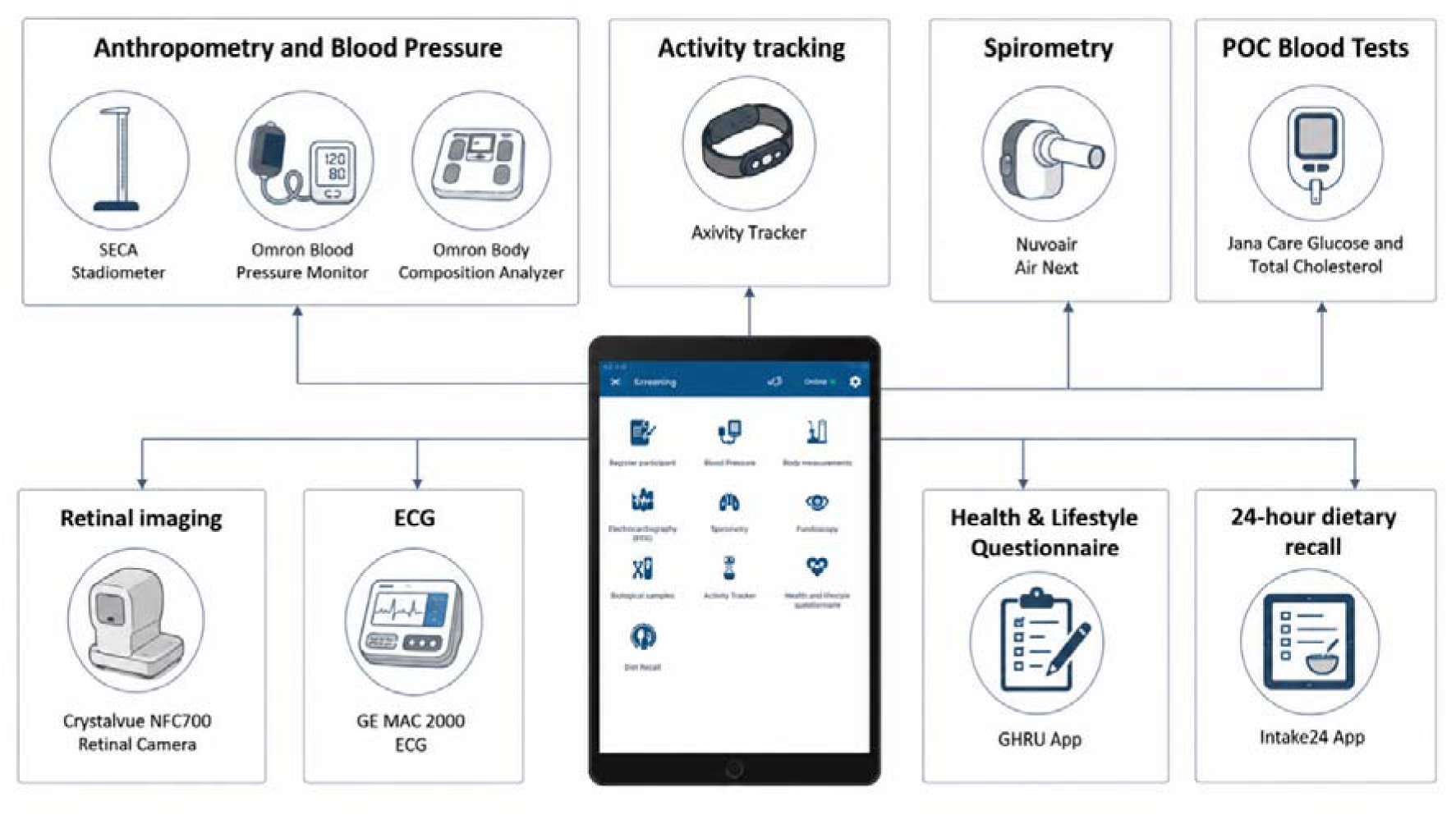
Summary of modular health assessment stations driven via the Android tablet application

#### Automated integrations with screening equipment

To enhance the data collection workflow and research dataset, data integrations were built where possible, between the platform and screening equipment in order to automate data capture into the database and reduce potential errors in data entry. Different integration approaches were required to cater to the range of data transfer options offered by equipment manufacturers (Figure 2). These applied a pragmatic and creative approach to solving the integration challenges. Where possible existing integration endpoints were chosen (such as ability to use in-built Wi-Fi and SFTP support on the ECG). For app integrations, the Android intent filter was the preferred method allowing data exchange between apps on the device. Whilst this requires both apps to be resident on the tablet, it enabled the team to avoid costly SDK development and harness the rich features of the native app to guide effective data collection (such as with the NuvoAir).

Examples of data integration approaches include transfer of full raw data in Extensible Markup Language (XML) format for ECG scans via automated file transfer protocol (FTP) transfer provided natively by the ECG machine and the use of tablet app-to-app data transfer (described above) to capture spirometry flow curve raw data in JavaScript Object Notation (JSON) format from measurements performed in the manufacturer’s app via custom application programming interfaces developed in collaboration with the manufacturer (**Figure 4**, panel A). This approach was chosen over an SDK integration as it meant all of the highly specialised User Interface for guiding a patient through a spirometry session were provided by the application (not within the screening platform).

**Figure 4.**
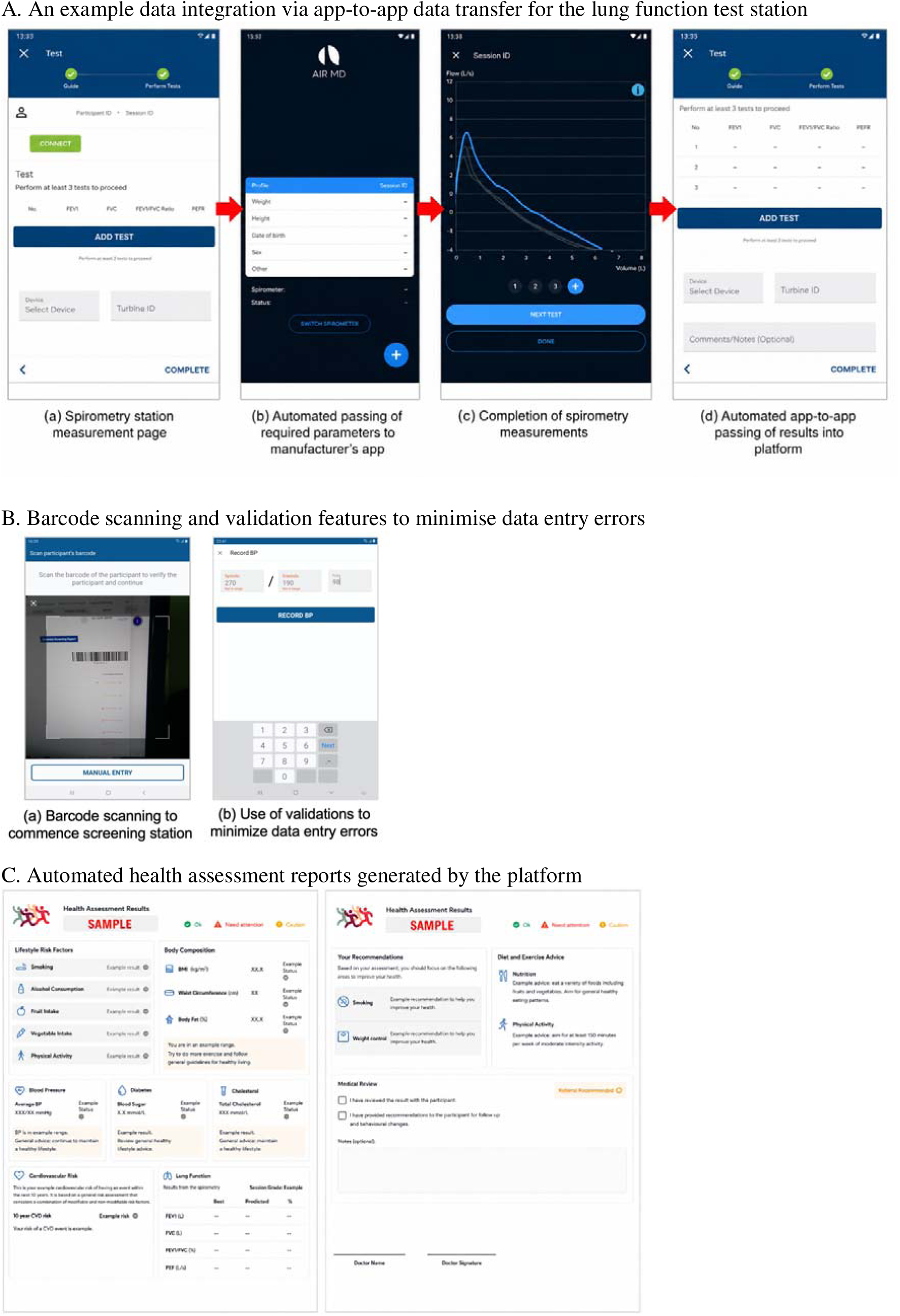
Illustration of selected functions

#### Use of barcode scanning and validations to minimise data entry errors

In order to reduce errors in data entry, barcode scanning functionality was built into the app to allow efficient entry of participant IDs at each station as shown in **Figure 4** panel B(a). Barcode scanning of participant IDs are used to initiate each station and verify the participants identity. A checksum digit was also incorporated to the participant ID and barcode to provide an additional layer of protection for data integrity. In addition, field validations were added for each data entry field to restrict input to valid ranges and prompt data collectors when a data entry error has been made as in **Figure 4** panel B(b).

#### Automated generation of a health assessment report for participants

In order to provide feedback to study participants on their health status, inputs captured during the health assessment are processed and presented as a report which is printed and provided to the participants (**Figure 4**, panel C). The report is generated through an integration with an API algorithm provided by the Open Health Algorithm Service (OHAS). Using the OHAS service, a custom algorithm based on the WHO Heart guidelines for generating recommendations based on WHO/ISH CVD risk stratification for each of the regions was published and validated. OHAS provides configurable algorithms to adapt the output for each setting [22], and processes health measurement inputs and provides health status assessments for a range of key metrics including body mass index, blood pressure and blood glucose. In addition, the algorithm calculates the participant’s CVD risk score according to the WHO HEARTS guideline and provides recommendations for lifestyle interventions for the participant. The OHA service also allows tailoring of recommendations content to fit the local context and language without requiring changes to the underlying algorithm. These recommendations were validated with each of the study teams to confirm suitability in each setting.

#### Administrator dashboards

A purpose-built administrator dashboard application accessible on a web browser on the local server was developed to help field team managers and study investigators with the management of the surveillance studies (**Figure 5**). Key functionalities include the ability to (i) track participant progress through the screening process; (ii) review data being collected in real time; (iii) generate participant health assessment reports; (iv) manage platform user access and permissions; and (v) register and manage study information such as screening site details and records of screening devices.

**Figure 5.**
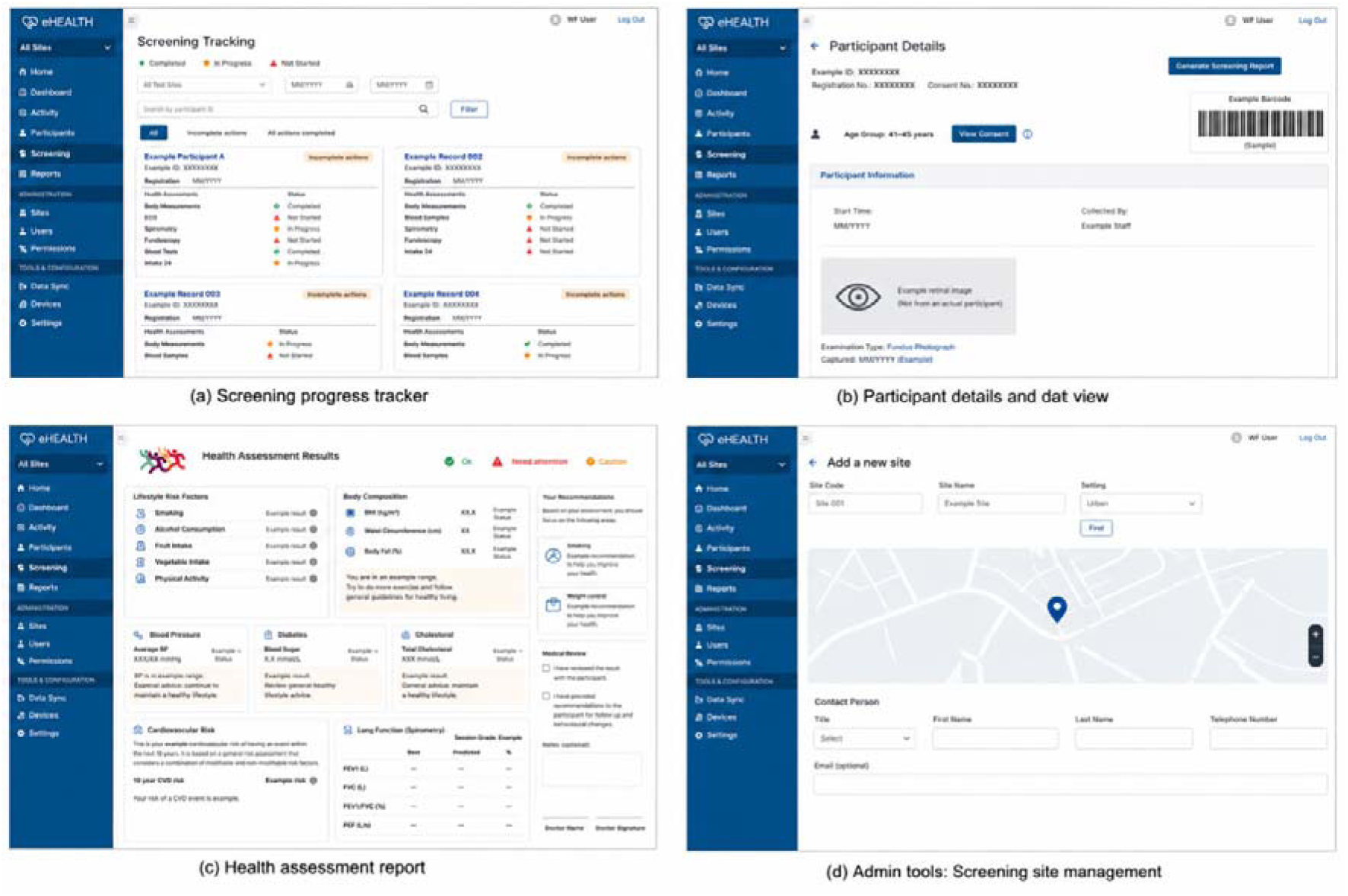
Key administrator dashboard features to support study management

#### Cloud data upload

To ensure captured data are secured, backed up and accessible by the study investigators, data on the local servers of each of the 12 screening teams are uploaded via an encrypted connection to region-specific cloud servers hosted by Google Cloud Platform (GCP). Once in the cloud, the data can be accessed by approved study investigators for analysis and quality control checks. The platform uses both in-built application-level authentication as well as GCP IAM controls to secure access to the data sources.

#### Monitoring and quality control tools

To maintain the pace of participant recruitment and quality of data, GCP tools for organising and visualising were used to create automated reports and asset sampling workflows that can be used to track team progress and data quality (**Figure 6**). These tools are used to identify potential issues in the data collection process for feedback to the respective teams for immediate rectification.

**Figure 6.**
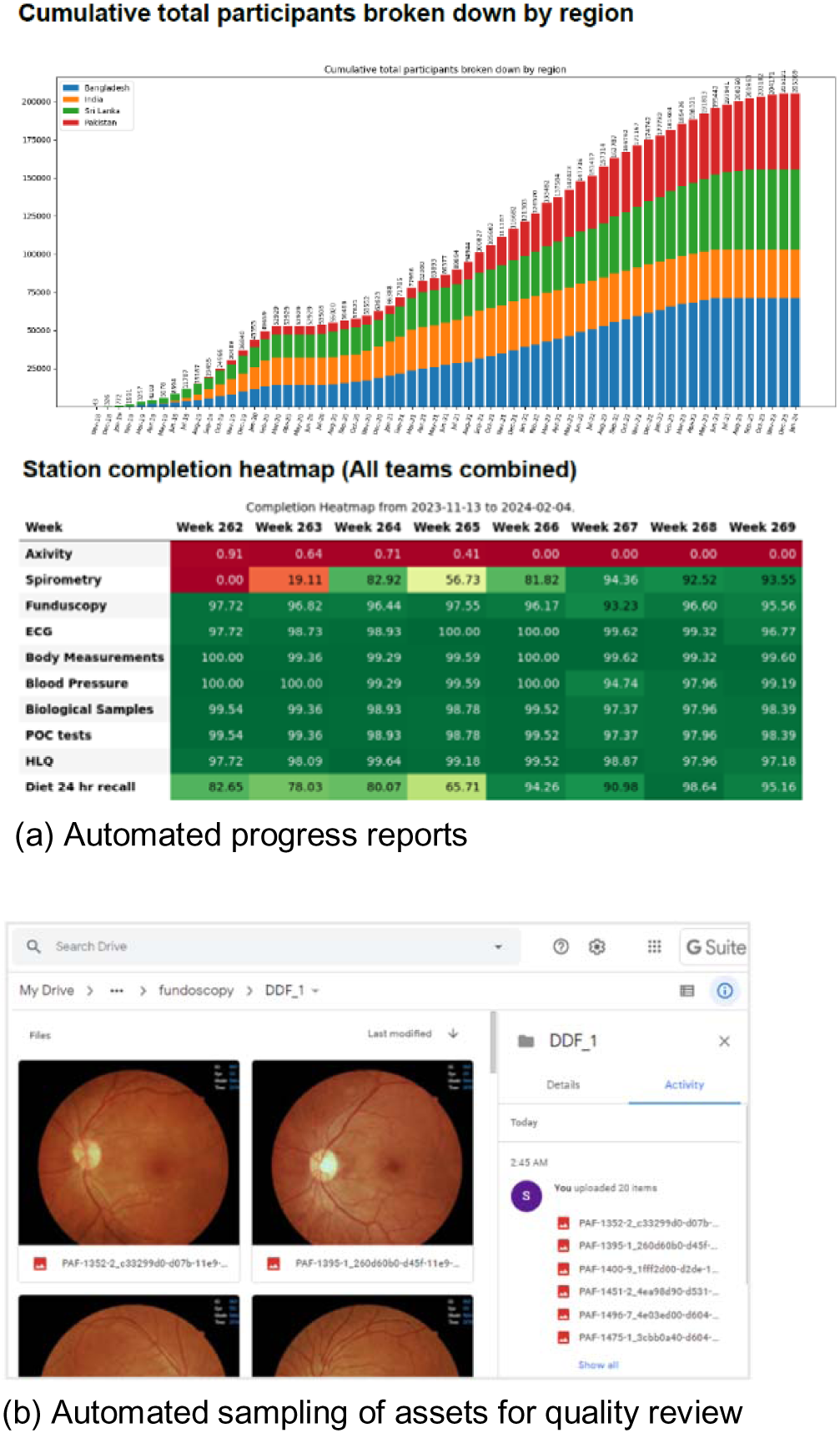
Cloud-based monitoring and quality control tools integrated into platform

### Platform deployment

In parallel with the platform development, a comprehensive deployment plan was prepared to ensure a smooth rollout of the surveillance study across multiple countries, focusing on consistency in health assessments and data collection protocols across all settings. The key elements of the deployment plan included a comprehensive manual of procedures, user training activities and materials, delivery of platform and screening equipment, post-deployment data quality assessments and ongoing platform support and maintenance. Once the major development tasks were complete, platform and screening equipment were procured and delivered to the field teams in each country. Based on experience from prior studies, comprehensive training was identified by the study investigators as critical to ensuring study field teams were well-oriented on the platform and screening procedures. Three phases of training were conducted for each team prior to launching in the field. The first phase involved a central 3-day training session for team managers conducted by the study technology leads. This was followed by local training for the field team members conducted by the team managers, and then a third phase of training coupled with in-field pilot deployments conducted by a study technology lead. All training phases consisted of platform orientation presentations, practical sessions involving platform set up and practice on performing all stations and ending with a session to discuss feedback and areas for improvement. In all teams, the launch of surveillance proceeded smoothly due to this comprehensive training process, with teams quickly ramping up participant throughput to between 25 and 50 participants screened per day within a week of launching. The training was crucial to ensuring the platform was deployed effectively in each setting. Post-deployment, weekly surveillance quality control reports are generated and disseminated via group calls and individual team discussions. Feedback received from field teams on platform functionality, maintenance and enhancements are discussed and passed on to the technical team for implementation. The study administrators also conducted periodic site visits to ensure protocols were being adhered to.

Development and deployment of the platform were achieved at modest cost relative to the scale of the study. The core software development effort for a custom platform, spanning the four user-centred design loops over approximately five months, and encompassing the tablet application, administrator dashboard, device integrations, cybersecurity assessments and cloud infrastructure, was completed for approximately USD 200,000. Ongoing cloud hosting, data storage and transfer through Google Cloud Platform amounted to approximately USD 24,000 per year, across the region-specific instances, a component partially offset by cloud credits provided by Google. Training and field deployment, comprising the three-phase training programme, development of the manual of procedures and training materials, and in-field pilot support across the 5 regions added approximately USD 20,000.

SAB baseline health assessment data collection for more than 205,536 participants from 564 surveillance sites across 5 regions was completed in January 2024. The data collected across the multiple screening locations was found to be consistent and of high quality. Independent assessment of a representative sample of asset files generated from the retinal imaging and physical activity stations show an average of >90% of the assets to be of high quality, consistently across the study regions. In addition, only a small number (<100 participants) of data entry errors outside of normal measurement ranges were identified during the analysis of the data, primarily due to human error.

## Discussion

In this paper, we describe the successful development of a robust and scalable health assessment data collection platform that can be deployed without the need for an internet connection. The platform leverages advances in mobile and medical device technology along with software integration capabilities to enable efficient and consistent data collection and the ability to review data quality in real-time. The user-centred design methodology employed to iteratively build the platform was critical in developing a platform that met the demanding requirements of the deployment settings, and satisfied study protocol requirements, while also providing a superior user experience for data collectors, all of which supports higher quality data collection. Testing of prototype applications and equipment in real-world settings provided critical usability insights and allowed platform designers to observe first-hand the challenges faced by field teams and implement solutions to address these challenges. An example of an issue observed was the occasional occurrence of tablet barcode scanners failing to detect barcodes, which led to implementing a manual option for participant ID entry. The use of quantitative assessments of platform usability via the SUS, coupled with qualitative interviews with different users to evaluate the platform enabled the software development team to prioritise their efforts on building the most relevant features. The increase of SUS scores through each iterative loop were valuable indicators for the progress of platform development to pilot and complete deployment phases.

The total cost of developing and deploying nHealth compares favourably with digital data collection systems reported in comparable multi-country and LMIC research settings. Most large field studies have adopted off-the-shelf, open-source toolkits such as Open Data Kit (ODK) and report only modest running costs, for example, the Strategic Typhoid Alliance across Africa and Asia (STRATAA) captured data from over 308,000 individuals across Malawi, Bangladesh and Nepal using an ODK-based system at an estimated variable cost of approximately USD 13,800 per site [23], while an ODK household survey in the Niger Delta was deployed for roughly USD 207 per survey round, less than half the cost of the paper-based equivalent [24]. Similar electronic systems for influenza surveillance in Kenya incurred higher start-up costs but lower total costs than paper over two years of operation [25], and the LINKS platform demonstrated that open-source, Android- and web-based tools can substantially reduce device acquisition and shipping costs for global health programmes [26]. Our one-off development cost of approximately USD 200,000 is higher than these figures because, unlike survey-configuration tools, no existing platform met the study’s overarching requirements for offline operation, automated integration with heterogeneous screening devices, and end-to-end surveillance workflow support, necessitating a bespoke build. Set against the scale of the study, however, the combined development, cloud and training investment equates to well under USD 2 per participant across the 205,536 individuals screened, a figure that is highly competitive given the breadth and depth of data captured (over 1,000 data points per participant).

Consistent with broader evidence that the cost-effectiveness of electronic versus paper-based capture is highly context-dependent [27], the principal cost drivers in our setting were the software engineering effort required to build custom integrations with each manufacturer’s equipment and the substantial investment in multi-phase, in-person training judged essential to consistent deployment across culturally and geographically diverse sites. Conversely, several deliberate design choices contained costs: the use of open-source frameworks (Laravel, PostgreSQL, Android) that avoided recurring licensing fees and kept maintenance expertise widely available; low-cost Android tablets; app-to-app integration methods such as the Android intent filter that avoided costly device-specific SDK development; a modular architecture replicated across all sites rather than rebuilt for each; and offline-first operation that removed any dependence on continuous connectivity. The resulting low marginal cost of adding each new site or country is an important consideration for the scalability of surveillance platforms in resource-constrained settings. As with any single-study costing, these figures reflect the specific labour markets, partnership arrangements and in-kind contributions of the SAB, including cloud credits and equipment support from Google, and may not transfer directly to other contexts.

Several digital platforms have been described for research data collection in comparable settings, but to our knowledge none combine the full set of capabilities offered by nHealth. Existing solutions tend to fall into two groups. The first comprises portable, offline-capable, open-source tools developed for field research in low-resource settings, such as the Open Data Kit–based electronic data capture system deployed across Malawi, Bangladesh and Nepal in the STRATAA typhoid surveillance programme [23], the REDCap Mobile Application designed for research in regions with intermittent connectivity [28], and the LINKS Android- and web-based system for neglected tropical disease programmes [26]. These systems are well-suited to portable deployment but are oriented primarily towards questionnaire and manual data entry and do not provide automated capture from a range of medical screening devices. The second group comprises richly device-integrated assessment platforms, exemplified by large prospective cohorts such as UK Biobank, which combined touchscreen questionnaires with integrated physical measurements and biological sampling [29]; however, these operate through fixed, well-resourced and reliably connected assessment centres rather than portable field configurations suited to remote low- and middle-income settings. nHealth is distinguished by sitting at the intersection of these approaches — offline-capable, portable and built on open-source frameworks, while also enabling automated, integrated capture from a wide variety of health screening equipment tailored for deployment across diverse LMIC settings. The rigour of our user-centred design process is reflected in the platform’s System Usability Scale score of 81.8, which exceeds published benchmarks for comparable field data collection applications, including the 74 reported for the offline data capture app used in the multicentre digiDEM Bayern registry, albeit assessed in a different setting [30].

This work has several limitations. First, usability was assessed with a small group of ten research assistants who, while experienced in field data collection, were relatively young and homogeneous and were drawn from a single setting; their feedback may not fully capture the diversity of experience, language and digital literacy among the field teams across all four countries, and the System Usability Scale, though widely validated, provides only a summary measure of perceived usability. Second, we did not conduct a formal comparative evaluation of the platform against paper-based collection or against existing off-the-shelf tools, so the efficiency and data-quality benefits we describe, although consistent with the broader literature, are not derived from a controlled comparison. Finally, the costing presented is an indicative, single-study estimate taken from a research-programme perspective; it excludes screening equipment and field-staff costs, relies on in-kind contributions specific to the South Asia Biobank, and reflects local labour markets and partnership arrangements, and therefore may not transfer directly to other settings.

Envisaged as a resource for use by the wider research community, the source code for the platform, which we have named ‘nHealth’, will be made available on a reasonable-request basis. Further development to extend the functionality of the platform for the SAB will include terminology mapping support more built-in tools for automated quality control assessments, over-reading functionality for expert review and grading of selected measurements (e.g. ECG scans), as well as a follow-up module to support longitudinal data collection as the study progresses from baseline to later phases.

## Conclusions

Advances in digital technologies have enabled the collection of consistent and high-quality epidemiological data in research studies of the scale and scope of the SAB, which were previously prohibitively costly and operationally challenging. We have shown in this work that modern software and connected devices integrated into a comprehensive data management platform using UCD principles, can enhance user experience and drive consistent and high-quality data collection in population health studies. To the best of our knowledge, this is the first platform that enables integrated capture of health assessment data from a wide variety of screening equipment that is tailored for deployment in a range of LMIC settings.

## Data Availability

All data produced in the present study are available upon reasonable request to the authors

## ACKNOWLEDGEMENT

All authors thank all the team members and all participants in the South Asia Biobank. The authors would also like to acknowledge Google, LLC for the provision of unrestricted gifts for purchases of screening equipment and credits for the use of Google Cloud Platform hosting and services.

## FUNDING

This research was funded by the National Institute for Health Research (NIHR) (16/136/68 & 132960) using UK aid from the UK Government to support global health research. The views expressed in this publication are those of the author(s) and not necessarily those of the NIHR or the UK Department of Health and Social Care. SB was supported by the UK Medical Research Council (MC_UU_12015/3 and MC_UU_00006/4).

## AUTHORS’ CONTRIBUTIONS

JC, IG and FH contributed to the conception and design of the study. IG, FH, JC, AG, MH, AK, KO, MKM and RMA worked on the development of the digital platform. IG, FH, WX, VR and AK contributed to the implementation of data collection and management. IG and WX implemented the analysis of the data and wrote the first draft of the manuscript. All authors contributed to the interpretation of the data and the revision of the manuscript.

## COMPETING INTERESTS

FH is an employee of Google DeepMind. Google, LLC provided unrestricted gifts for the purchase of screening equipment and credits for the use of Google Cloud Platform hosting and services, as noted in the Acknowledgements. The remaining authors declare no competing interests. The funders and Google had no role in the study design, data collection and analysis, decision to publish, or preparation of the manuscript.

## DATA AVAILABILITY

Reports and major results of the South Asia Biobank (SAB) are released regularly on the SAB website (https://www.ghru-southasia.org/). Owing to cybersecurity considerations, the platform source code is not publicly available but can be made available on reasonable request, subject to a data/code-sharing agreement. Requests should be directed to Professor John C. Chambers.

## Abbreviations

CVD: cardiovascular disease
FTP: file transfer protocol
GCP: Google Cloud Platform
JSON: JavaScript Object Notation
LMIC: low and middle income countries
NCD: non-communicable diseases
OHA: Open Health Algorithms
SAB: South Asia Biobank
SUS: System Usability Scale
T2DM: type-2 diabetes mellitus
UCD: user-centred design
XML: Extensible Markup Language

---

**Supplementary Figure 1:**
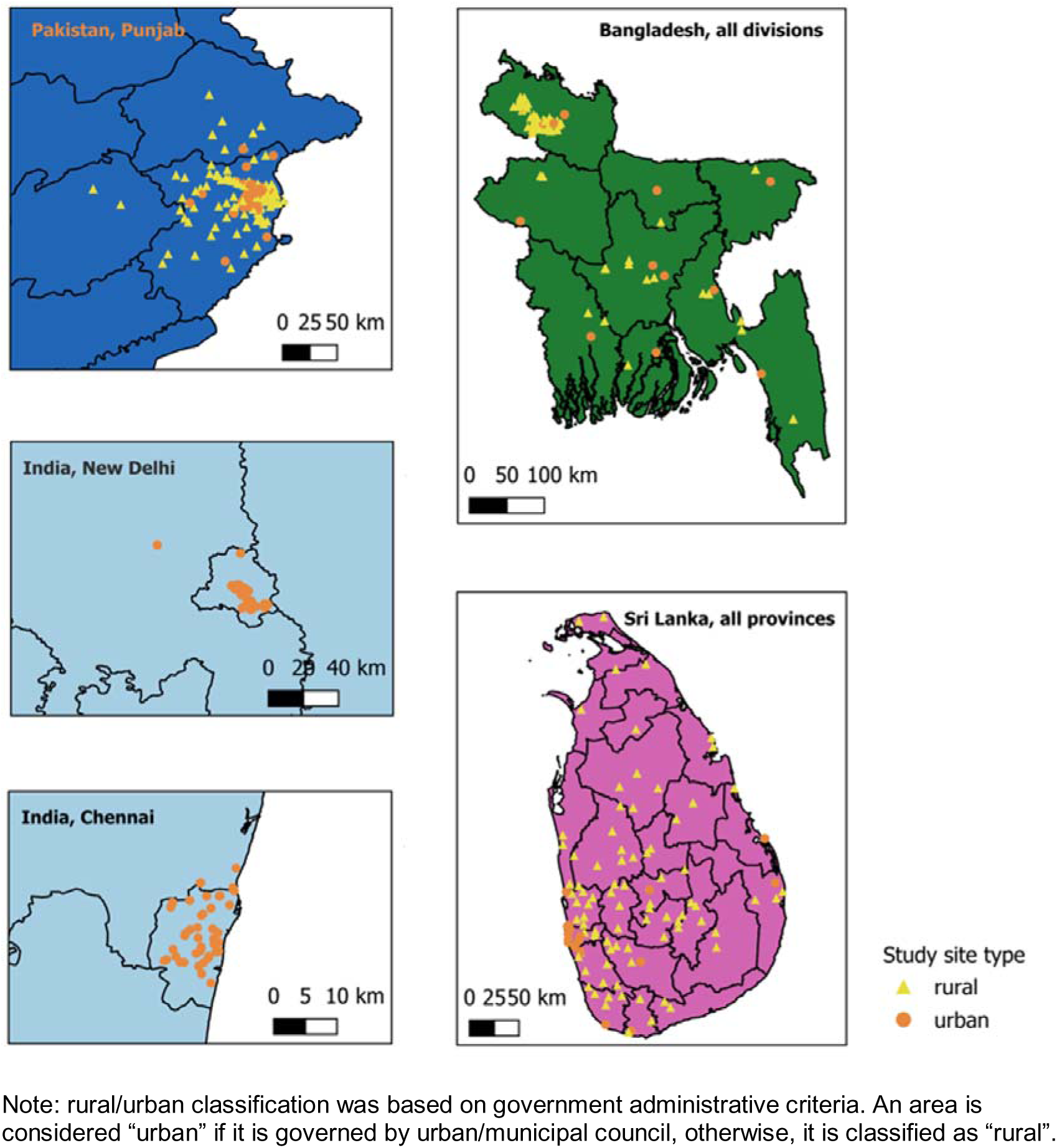
Map of the Locations of the South Asia Biobank surveillance sites

## Notes

### Competing Interest Statement

The authors have declared no competing interest.

### Author Declarations

The study is IRB approved in all countries (reference: Imperial College London Research Ethics Committee, United Kingdom: 18IC4698, Sri Lanka: EC-18-094; Pakistan: IRB/2018/424/SIMS; India North: RS/MSSH/DDF/SKT-2/IEC/ENDO/19-35; India South: IRB00002640; Bangladesh: BMRC/NREC/2019-2022/126), and written consent using translations to local languages and supported by infographics and videos, was obtained from all participants.

